# Gut Bacterial Microbiome Profiles Associated with Colorectal Cancer Risk: A Systematic Review and Meta-Analysis

**DOI:** 10.1101/2021.06.05.21258404

**Authors:** Christian A Russ, Nicholas A Zertalis, Veronica Nanton

## Abstract

**Objective:** Recent studies have shown a potential link between gut microbiome and colorectal cancer (CRC). Initially, a wide array of research into this topic was discovered from the past decade, illustrating a keen interest in the potential causal relationship between the gut microbiome and CRC. However, the cancer research community is lacking a summarised systematic review of this kind which aims to explore the evidence linking the human gut microbiome to risk of CRC.

**Design:** This systematic review was carried out with two independent reviewers assessing the database outcomes from Medline and EMBASE during May 2020. A meta-analysis was undertaken studying the link between *Helicobacter pylori* and CRC; processed through Stata.

**Results:** 31 papers were included in the systematic review, followed by 12 for the meta-analysis. From these papers, *Fusobacterium* and *Bacteroides* were reported most frequently as enriched in CRC versus control. The meta-analysis showed an Odds Ratio of 1.49 (95% CI 1.19 – 1.86), including a total of 20,001 events. This meta-analysis concluded that *H. pylori* infection significantly increases the risk of CRC, albeit with evidence of publication bias.

**Conclusions:** Bacteria have been discovered to increase the risk of CRC, however a definitive causal relationship cannot be concluded or excluded using case-control studies. To fully understand the potential link of the bacteria listed, alterations in research design and execution are required. The assessment found a need for a large-scale cohort study conducted over a significant period of time to thoroughly evaluate the potential relationship between gut microbiome and CRC risk.

Significance of study
**What is already known on this subject?**

➢ Roughly 10% of all cancer deaths in the UK are attributed to colorectal cancer (CRC), with CRC being the third most common cancer worldwide.
➢ The risk of developing CRC has been closely linked to the composition of the gut microbiome
➢ *H. pylori* is a known causative agent of gastric cancer

**What are the new findings?**

➢ To the best of the researchers’ knowledge, this is the first systematic review of this type conducted into this topic, investigating the genera/species of bacteria in the human gut microbiome and the risk of CRC.
➢ This systematic review found a strong association between *Fusobacterium* and *Bacteroides*, amongst other species, and CRC.
➢ The meta-analysis found a significant link between *H. pylori* infection and increased risk of CRC.

**How might this impact on clinical practice in the foreseeable future?**

➢ This systematic review provides potentially actionable evidence in the personalised management of patients to reduce their risk of CRC.
➢ This review has highlighted the need for a large population prospective cohort study, with standardised sampling methods.
➢ The meta-analysis reinforces the importance of *H. pylori* testing and eradication in those deemed at high risk of CRC

## INTRODUCTION

Colorectal cancer (CRC) is the third most common cancer worldwide,[1] accounting for 10% of all cancer deaths in the UK during the period of 2015-17.[2] The global burden of colorectal cancer is expected to substantially increase in the next two decades, as a result of the widening adoption of a western lifestyle.[3] Cancer Research UK estimates that 54% of CRC cases are preventable,[4] with many studies looking into the impact that lifestyle and other preventable factors can have on CRC risk, such as red meat consumption,[5] fibre intake,[6] and obesity.[7]

Recently, risk of developing CRC has been closely linked to the composition of the gut microbiome, with many papers stating evidence for and against certain commensal species normally found in the human gastrointestinal (GI) tract. It has been suggested that an understanding of this gut flora composition offers potential in terms of the identification of biomarkers,[8] and associated risk factors for early CRC. This may have an important impact on the future personalised management of patients, potentially improving prognoses.

The aim of this systematic review was to determine what genera/species of bacteria in the human gut microbiome are significantly linked to increased or decreased risk of colorectal cancers, through the evaluation of papers that have been published on this subject. The systematic review presented here summarises the bacterial taxa associated with altered risk of CRC, in line with this paper’s research question and the PRISMA statement.[9]

## METHODS

### Data sources & search strategy

This systematic review was reported in line with the Preferred Reporting Items for Systematic Reviews and Meta-Analyses (PRISMA) statement. The methods used were agreed by the authors in advance:

- Develop research question
- Identification of papers
- Screening
- Critical Appraisal
- Data extraction
- Narrative synthesis
- Meta-analysis

Studies that reported the association between CRC and gut microbiome were gathered from Medline and EMBASE, with the searches adapted to utilise relevant subheadings on each database. The following variations of keywords and MeSH terms were used: colorectal, cancer, neoplasm, tumour, malignancy, carcinoma, bacteria, microbiome, gastrointestinal, colonic, faecal, gut, dysbiosis. For the full unabridged search strategy please see Supplemental Material S1.

### Selection criteria

In accordance with the PICO (population, intervention, controls and outcomes) proforma, the following criteria were used for the search and selection of papers for inclusion and exclusion for the review (see Table 1). Both reviewers agreed to base the exclusion criteria around factors that may alter the natural composition of the gut microbiome found in humans, such as underlying conditions or medication exposure, which may inadvertently impact the composition of species. For example, a twin study by Willing et al.[10] in 2010 showed a significant difference in the gut microbiome of inflammatory bowel disease sufferers when compared with their healthy twin, suggesting a potential for underlying conditions to alter the host microbiome.

**Table 1.** inclusion and exclusion criteria

| Inclusion criteria | Exclusion criteria |
| --- | --- |
| • CRC | • Other cancers (gastric, small intestine etc) |
| • Human participants | • Studies in non-humans |
| • Case-control study or Cohort study or Cross-sectional | • Other study types and publication types – no reviews |
| • Adenoma & Adenocarcinoma or only Adenocarcinoma | • Adenoma only and/or no healthy control groups |
| • Published literature only | • Unpublished literature |
| • Available electronically | • Not available electronically |
| • No significant underlying conditions | • Underlying condition in participants or medication exposure (antibiotics, PPI, chemotherapy) |
| • Specify bacteria in microbiome | • Microbiome metabolites linked to CRC |
| • Must directly link microbiome species associated to CRC | • Microbiome linked to CRC prognosis/clinical outcome/treatment efficacy/procedure evaluation |
| • Bacterial differences | • Viral or fungal focus |
| • Named bacteria or a known origin | • Ethnic and socioeconomic microbiome linkage |
| • Taxonomic level reported | • Dietary effect on microbiome/CRC |
| • English language only | • Probiotics to treat complications in CRC patients |
| • Full text available | • CRC associated bacteraemia/endocarditis |
| • Raw data for meta-analysis | • Post-op surgical site infections |
| • Original results | • Genetic involvement/expression in the progression of CRC |

The literature search, selection and review were performed by two independent reviewers. Papers were removed from the selection if both agreed to exclude them at the various screening levels (title & abstract and full article eligibility), or if duplicates were found. In instances of disagreement, a resolution was reached via discussion between the two study team members. The overarching objective of the reviewers at this stage was to focus on the two absolutes, that being groups that are minimally affected by possible gut altering effects outside of the parameters measured.

### Data extraction

Data extraction was carried out independently by the two reviewers following the full article assessment and were documented on an original electronic data extraction table. Extracted data included: Author name, publication year, population location, population size, detection method, taxa enriched in CRC, taxa enriched in control. The same data was extracted for the meta-analysis, with the number of *Helicobacter pylori* positive and negative patients documented for CRC and healthy groups replacing taxa documentation.

### Critical appraisal

Two reviewers performed a critical appraisal of selected articles. The relevant Critical Appraisal Skills Programme (CASP) checklist was used for cohort and case-control studies. The appropriate Newcastle-Ottawa Checklist was used for cross-sectional studies.

### Main outcomes measured

Significantly raised (P-value <0.05) levels of bacterial taxa in patients with and without CRC were recorded in the data extraction table. The meta-analysis was specific to *Helicobacter pylori* presence in patients with and without CRC.

### Meta-Analysis

During the screening and eligibility phases of this review, it became apparent that several of the articles generated studied the association between *Helicobacter pylori* and CRC. To further infer significance and verify this link, a decision was made to perform a meta-analysis on the eligible papers generated from the documented search strategy. To perform the meta-analysis, StataSE 16.1 for Mac (StataCorp, Texas, USA) was used.

Significance of association was measured using Odds Ratio (OR) and 95% confidence interval (95% CI). Random-effects model was chosen to analyse the data due to the differences amongst studies, namely in study design. Funnel-plot and contour enhanced funnel-plots were generated to test for publication bias, alongside Egger test and trim-and-fill.

## RESULTS

A total of 1,058 abstracts were screened from Medline and EMBASE; the final count of articles included for this systematic review and meta-analysis were 31 and 12 respectively. The process and numbers are documented in Figure 1.

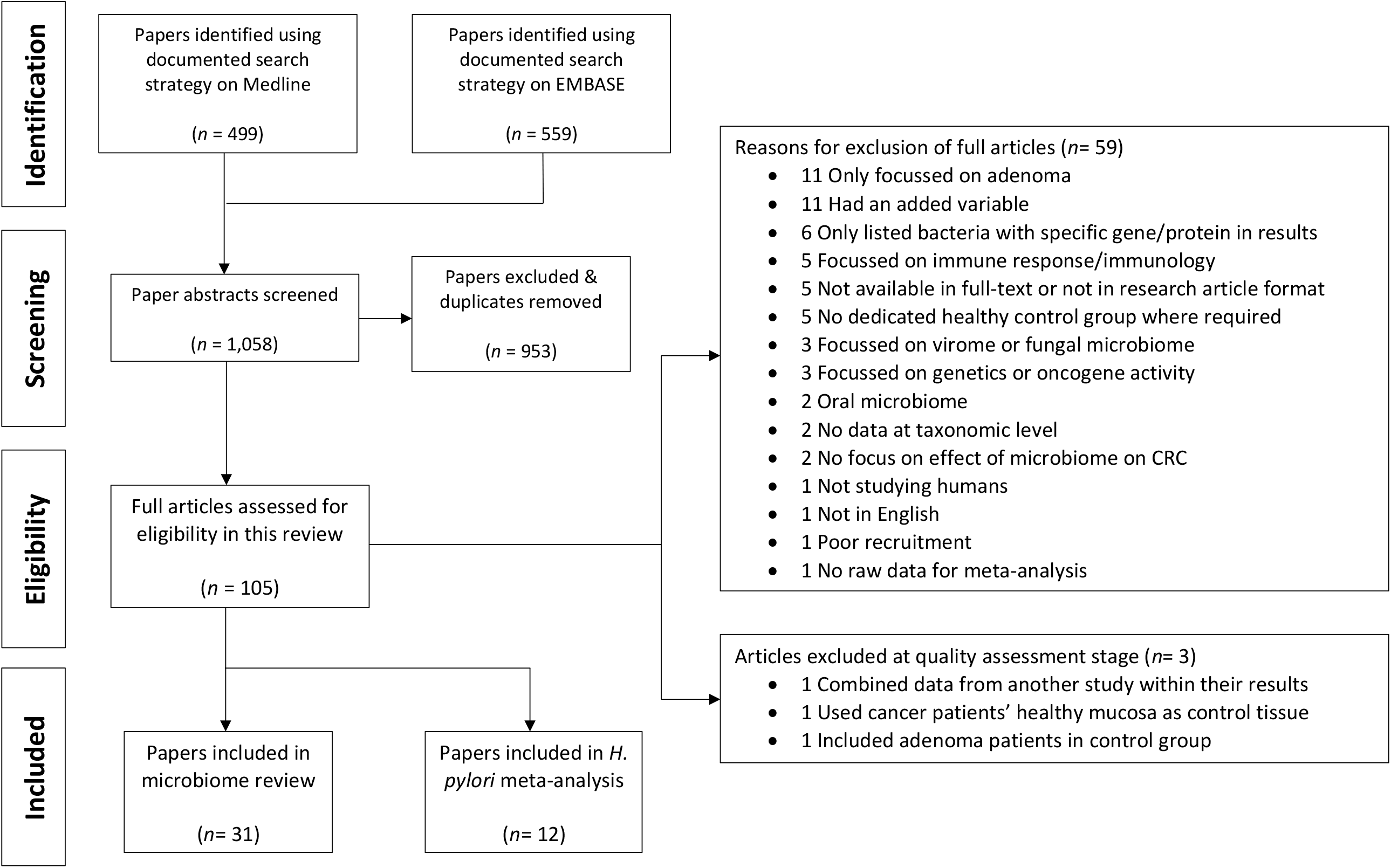

### Systematic Review

#### Study characteristics

A total of 30 out of the 31 articles included in the systematic review were case-control in design, and one a Cohort Study.[11] The studies recruited patients from a total of 20 countries, of which 7 studies were from China, 3 from USA, 3 Japanese, 2 Israeli and 2 Iranian. The median year of publication for all articles analysed was 2016, illustrating a recent increase in evidence and interest in this subject.

Many of the articles included for analysis had methods enabling the detection of a vast number of different taxa. Of the 31 studies included, 21 were set up in this way. The remaining 10 tested for specific species/genera of bacteria. The number of studies able to detect species within each genus can be found in Table 2. Please see Supplemental Information S2 for full details of the studies specific to certain bacteria.

**Table 2.** Number of papers with methodology specific to the detection of each genus

| Genus | Eligible Papers |
| --- | --- |
| ***Fusobacterium*** | 24 |
| ***Bacteroides*** | 23 |
| ***Escherichia*** | 23 |
| ***Streptococcus*** | 23 |
| ***Enterococcus*** | 22 |
| ***Clostridium*** | 21 |
| ***Roseburia*** | 21 |
| **All Other Genera** | 20 |

#### Quality assessment

The table summarising the outcome of all CASP checklists can be found in Supplemental Material S3. Three articles were excluded at the quality assessment stage, reasons for exclusion shown in Figure 1.

#### Significantly enriched genera in CRC and controls

Of all the taxonomic levels in the included papers, genus was most widely reported as being significantly raised in either group. Therefore, it was decided that genus and species level should be predominantly reported to provide more specific results. Significance was determined from the P-value tests used in each paper, recording any species or genus whose P-value was <0.05 in either group. The full data extraction table can be found in Supplemental Material S4. A total of 76 genera were either recorded as being significant, or a species within that genus was independently significant. When compared, 58 of those genera were recorded as enriched in CRC patients in at least one study, compared with 35 in controls. The frequency with which a genus or species was significant is recorded in Table 3.

**Table 3.**
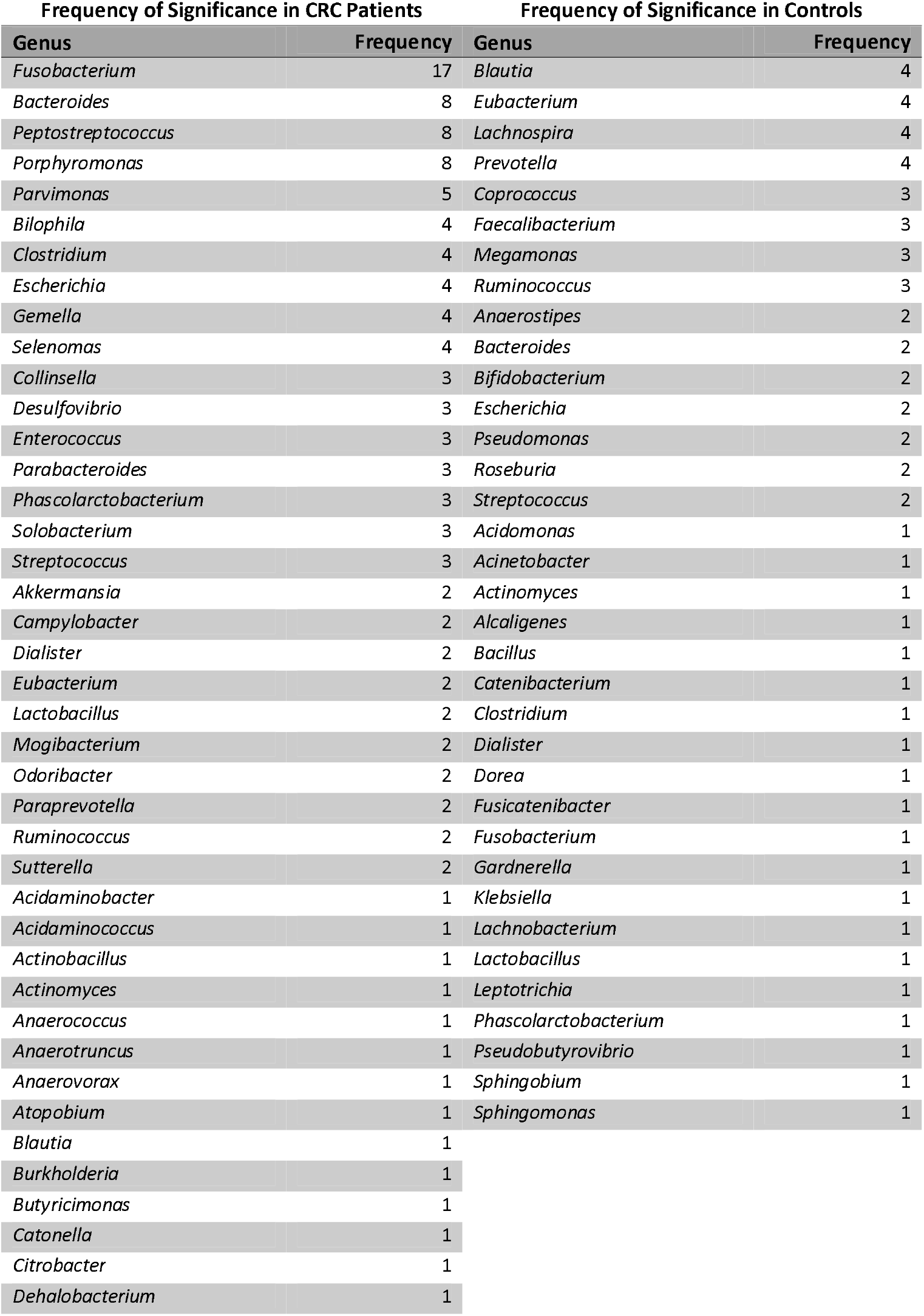
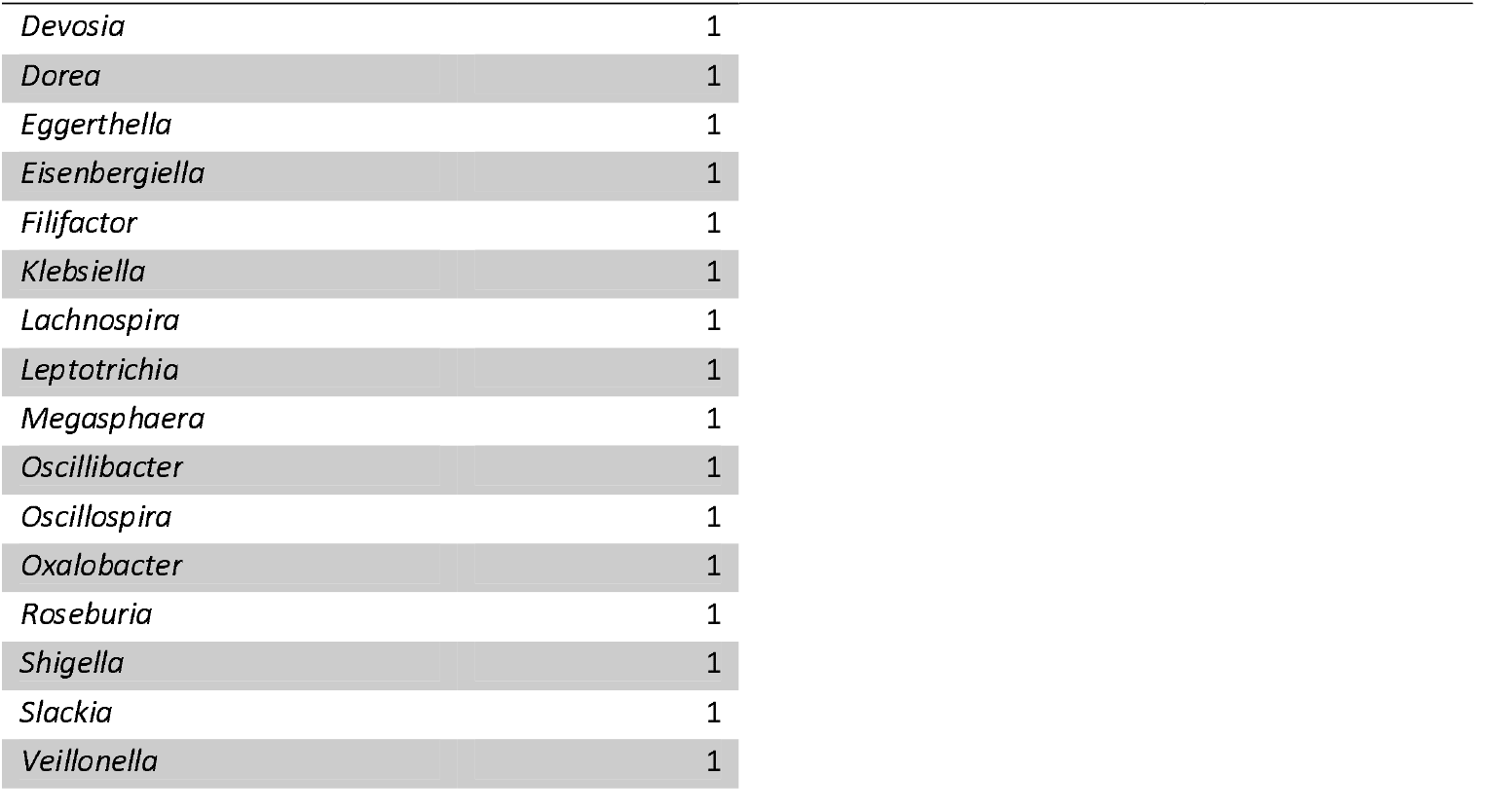
Number of articles in which a genus or species within a genus was statistically enriched

*Fusobacterium*, or a species within this genus, was recorded as statistically enriched in CRC patients in 70.8% of all studies powered to detect it. Within these studies, the whole genus was significantly enriched in a total of 11 papers, with individual *Fusobacterium* species enriched in 9 (1 of these in controls). *Peptostreptococcus* & *Porphyromonas* were raised in 40% of eligible studies, *Bacteroides* 33.3% and *Parvimonas* in 25% of studies. The median frequency of significance for all genera significantly enriched in the CRC group was 1 (IQR 2).

In contrast, *Blautia, Eubacterium, Lachnospira* and *Prevotella* were significant in 20% of eligible papers in control groups. The median frequency of significance within the control groups was 1 (IQR 1).

#### Significantly enriched species in CRC and controls

Of the *Fusobacterium* genus, *F. nucleatum* was the most commonly raised species amongst CRC patients in the papers analysed, statistically enriched in 7. Only 2 individual species were identified as significantly enriched in this group, *F. nucleatum* and *F. varium*, with the latter only enriched in a single study.[12] A third species, *F. peridonticum*, was recorded as significantly enriched in controls only once, in the same article by Saito et al.[12]

*Bacteroides fragilis* was significantly enriched among CRC patients in 5 studies, 2 of these looking specifically for this species. A total of 8 other *Bacteroides* species were recorded as enriched in CRC and 4 in controls, however each was only significant once. A summary of the species reported as significantly enriched in 2 or more papers is provided in Table 4.

**Table 4.** Number of papers reporting a species as significantly enriched

| Frequency of Significance in CRC |  | Frequency of Significance in Controls |  |
| --- | --- | --- | --- |
| Species | Frequency | Species | Frequency |
| *Fusobacterium nucleatum* | 7 | *Prevotella copri* | 3 |
| *Bacteroides fragilis* | 5 | *Prevotella stercorea* | 2 |
| *Gemella morbillorum* | 3 | *Eubacterium eligens* | 2 |
| *Parvimonas micra* | 3 |  |  |
| *Peptostreptococcus stomatis* | 3 |  |  |
| *Solobacterium moorei* | 3 |  |  |
| *Enterococcus faecalis* | 3 |  |  |
| *Escherichia coli* | 3 |  |  |
| *Bilophila wadsworthia* | 2 |  |  |
| *Collinsella aerofaciens* | 2 |  |  |
| *Clostridium symbiosum* | 2 |  |  |
| Enterotoxigenic *B. fragilis* | 2 |  |  |

Both *Enterococcus faecalis* and *Escherichia coli* were significantly enriched in CRC patients in 3 studies, however 2 of these came from papers that were looking specifically for those bacteria. *Streptococcus bovis* was specifically tested for in 3 studies, although only one of these studies returned a statistically significant enrichment in CRC patients compared to controls.

### Meta-Analysis

This paper’s meta-analysis was exclusively looking at the association between *H. pylori* infection and the associated risk of CRC. A total of 12 papers; 7 case-control, 2 cohort and 3 cross-sectional, were identified from the search strategy and eligibility criteria used for the systematic review. The Forest plot (Figure 3) is split by study design for transparency with an overall odds ratio. A full table of the study characteristics of those papers included in this meta-analysis can be found in Table 5.

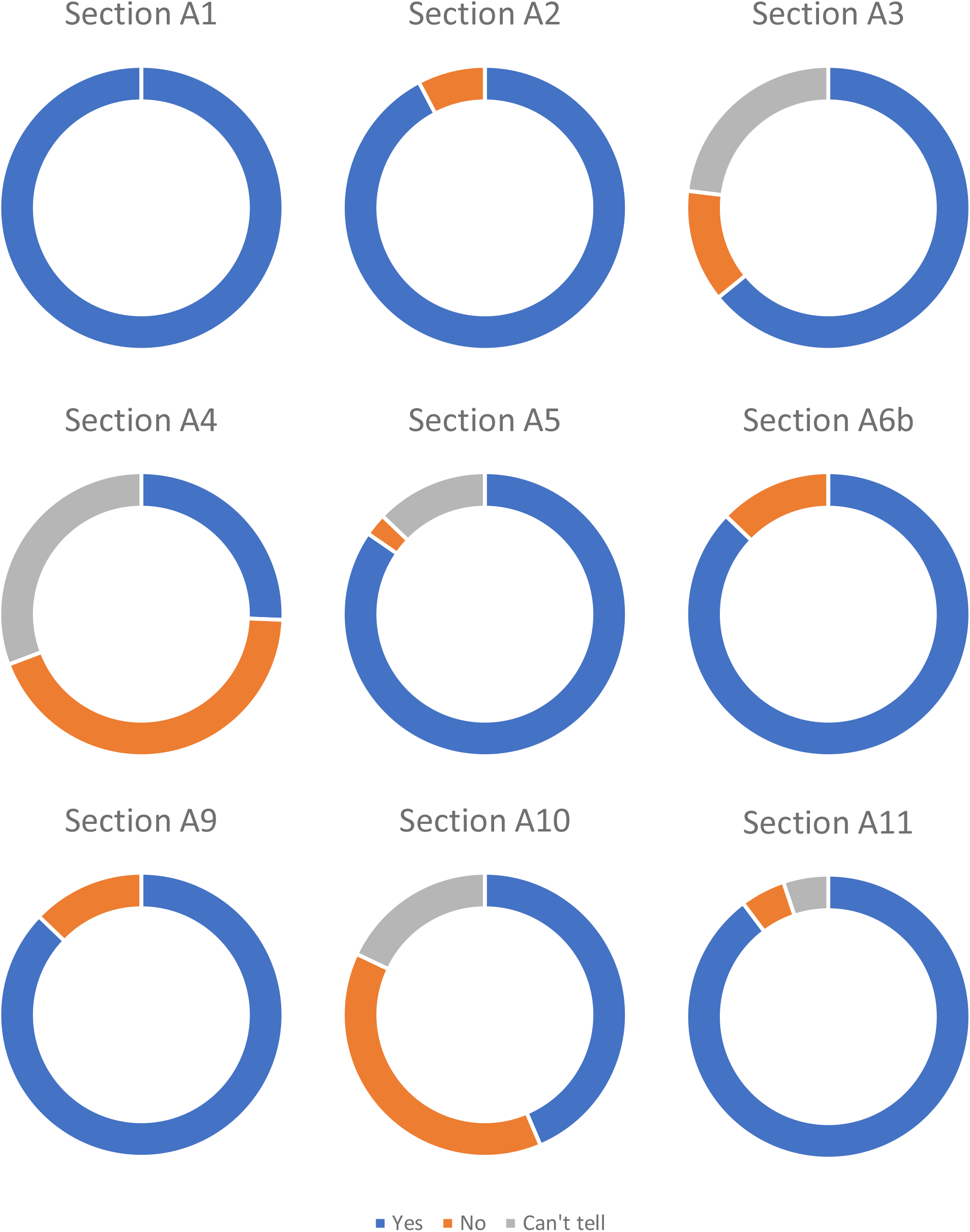

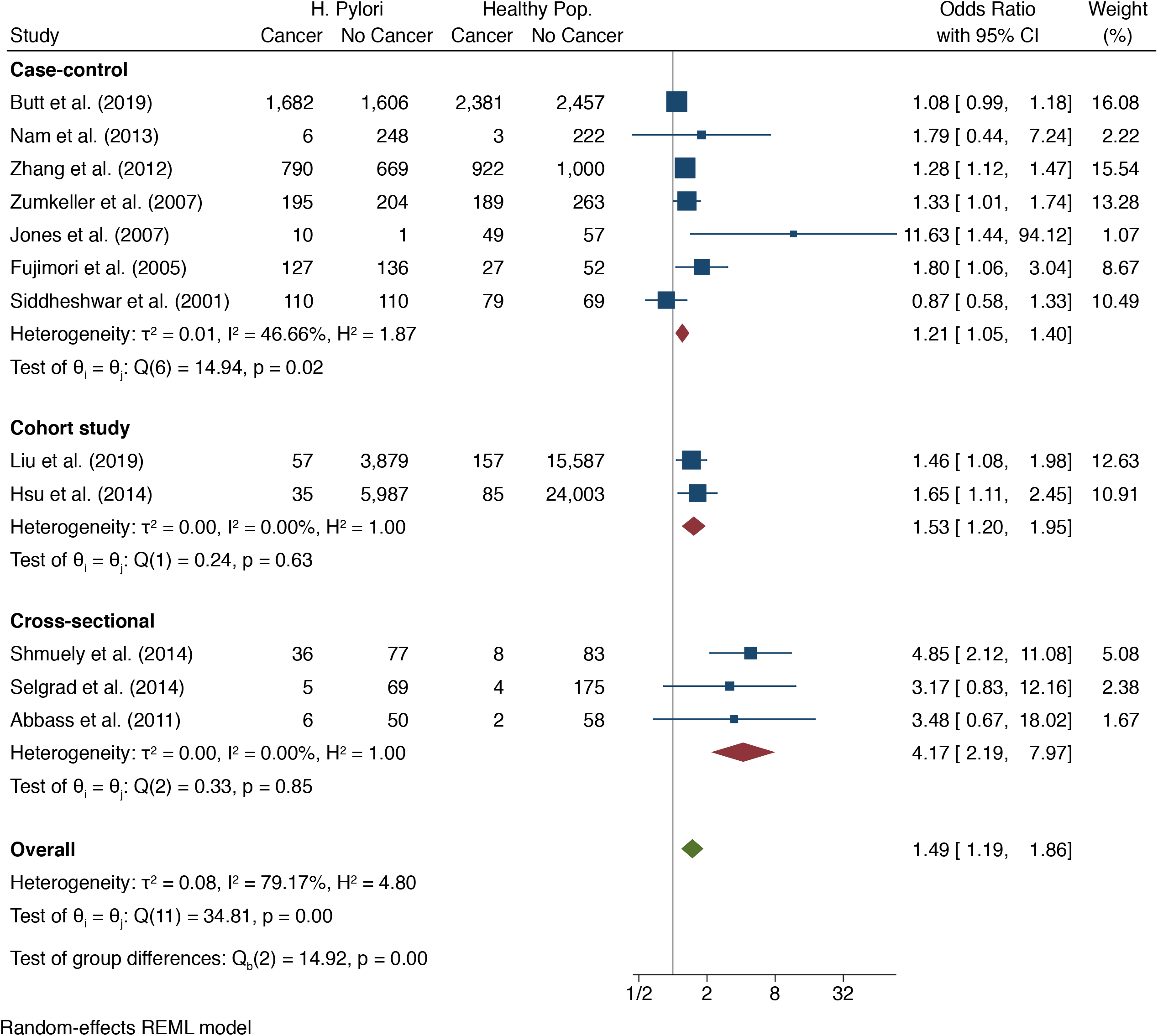

**Table 5.**
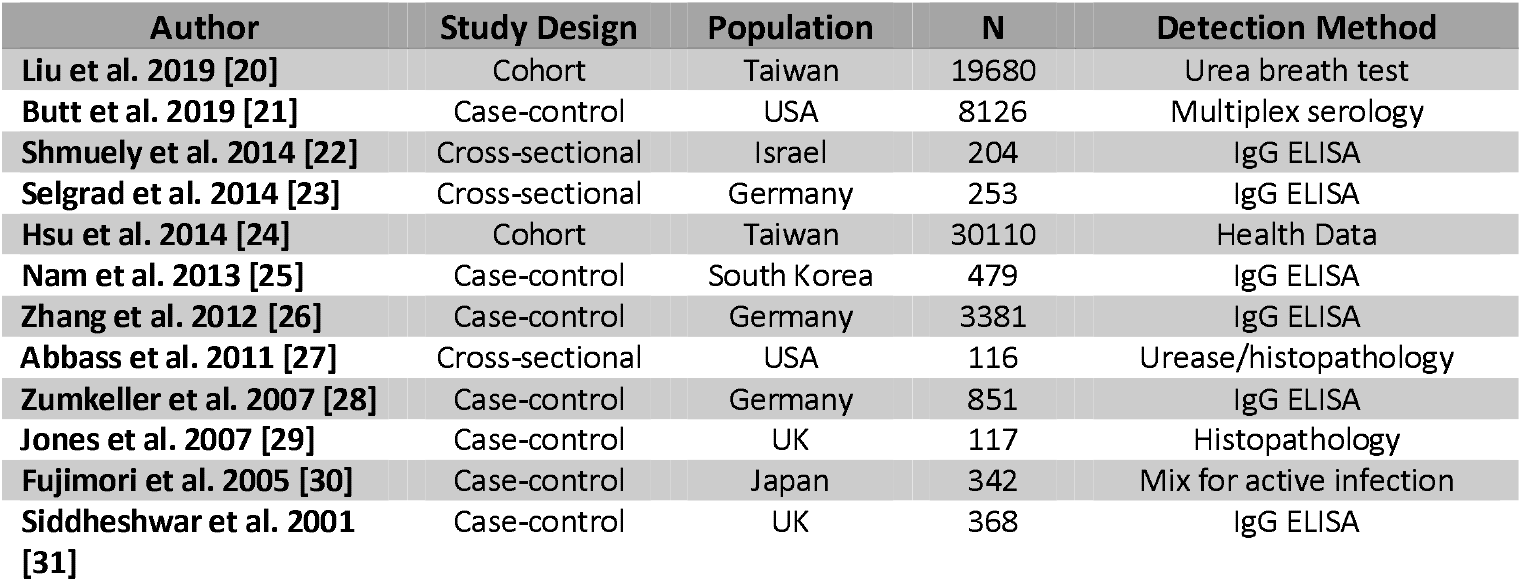
H.pylori included study characteristics

This meta-analysis found an OR of 1.49 (95% CI 1.19 – 1.86), showing a statistically significant link between *H. pylori* infection and increased risk of CRC (Figure 3). Upon exploration of publication bias, significant risk was found. Funnel plots proved to be asymmetrical and Egger test gave a *p*-value of 0.0001, showing evidence of bias. When corrected for with trim-and-fill, the calculated OR came to 1.26 (95% CI 0.93-1.71).

An I^2^ result of 79.17% shows substantial heterogeneity between studies, largely arising from the case-control group. The sub-analysis by study design shows a statistically significant link with each design, the largest effect being seen amongst cross-sectional studies. This meta-analysis included data of 20,001 events, out of a total of 64,027 participants.

## DISCUSSION

### Systematic Review

#### Results

Some bacterial genera were featured amongst studies at a higher frequency than others. *Fusobacterium* was documented by far the most at 17, reported as significantly enriched in CRC patients more than twice as often as *Bacteroides, Peptostreptococcus* and *Porphyromonas*, the next most common at 8. On the contrary, the most featured control group genera were only significant in 4 studies, with 16 of 31 studies reporting any significant enrichment in this group.

Table 3 illustrates a high disparity amongst studies, in the reporting of taxa significantly associated with CRC and controls. Of the 58 significant genera in CRC patients, 30 of these were significant in only 1 paper. Similarly, 20 of the 35 significant genera in the controls were associated in one paper. Additional affiliations can be seen at species level, as 57 different species were found to be significant in CRC patients in at least one study. The most featured of these was *Fusobacterium nucleatum* at 7, then *Bacteroides fragilis* at 5 (7 if including enterotoxigenic strains). Frequency of replication of these results, however, was low. Only 13 of the 57 species was featured more than once between papers, as can be seen in Table 4. This replication frequency was even lower for species enriched in controls, with only 3 of the 40 species across articles being replicated more than once.

The research included in this systematic review originated from a wide variety of countries. Papers have found differences in the microbiome between patients with varying diets,[13, 14] which could predispose participants with certain diets to already high levels of specific bacteria. Thus, this disparity may, in part, be due to the variations in diet between cultures.

#### Limitations of included studies

Recruitment methods were a limitation of many studies included in this review. Controls in most of these studies were not recruited effectively (please see CASP results of Q4 in Supplemental Material S3 for such papers). Most studies recruited from colonoscopy waiting lists, meaning that controls were presenting with colorectal symptoms warranting further investigation. Thus, the conclusion cannot be drawn that these are truly ‘healthy’ controls, and the control microbiome may have been altered as a result of the symptoms. Cases, however, were often recruited in an acceptable way, predominantly due to their diagnosis.

Papers also failed to consider the presence or absence of blood in the stools of the patients being studied. A recent article by Chenard et al.[15] showed significant differences between the gut microbiome of those with and without blood present in stools. Specifically, *Bacteroides uniformis, Clostridium symbiosum* and *Collinsella aerofaciens* were found to be significantly enriched in participants with bloody stools; all species found to be significantly enriched in CRC patients in this systematic review. The *Bacteroides* genus was also significantly enriched in this group. Conversely, *Faecalibacterium prausnitzii, Prevotella copri* and *Roseburia faecis* were enriched in the controls described by Chenard et al., as well as the controls in this systematic review. As blood in the stool is considered a key sign of CRC,[16] this could explain some of the disparities between the microbiome profiles of CRC and healthy individuals.

Many articles included in this review hypothesised that the microbiome has a causative role in CRC carcinogenesis. Causation, however, cannot be inferred through the study designs adopted. Case-control designs, as adopted by all but one study, cannot determine whether CRC is caused by the microbiome profile detected, or whether this profile arises due to environmental changes caused by CRC, due to testing at a set-point as opposed to over time. It is of no surprise that case-control design was used in the majority of studies; a causal relationship can often be inferred using these methods. However, in the case of studying the potential role of the microbiome in carcinogenesis, many factors may increase or decrease species abundance at a set point in time. Therefore, concluding that the enrichments are purely as a result of involvement in CRC carcinogenesis risks oversimplifying a complex array of interactions.

Microbiome sampling differed between studies. Included in this review, Chen et al.[17] performed a mix of rectal swabs, faecal sampling and biopsy for microbiome detection and found significant differences between each. Therefore, a truly representative microbiome profile may not have been obtained in many of the studies that used only 1 detection method.

#### Limitations of this Systematic Review

The large number of countries and thus potential difference in microbiome related to diet was not thoroughly evaluated. This paper’s focus on genus and species taxonomic levels may also be seen as overly specific, when a more thorough analysis of all taxonomic levels implicated in CRC may provide a broader consensus. As this systematic review excluded studies not written in English, this decision may have excluded papers that could have enlightened further on this subject.

### Meta-Analysis

#### Results

As can be seen in Figure 3, The OR of 1.49 (95% CI 1.19 – 1.86) shows a statistically significant link between *H. pylori* infection and CRC. However, this meta-analysis found considerable heterogeneity, which must be considered when inferring clinical significance. As part of the analysis, a funnel plot (Figure 4), contour-enhanced funnel plot, Egger test and trim-and-fill were run on the extracted data to search for evidence of publication bias. The latter can be seen in Supplemental Material S5, S6 and S7 respectively.

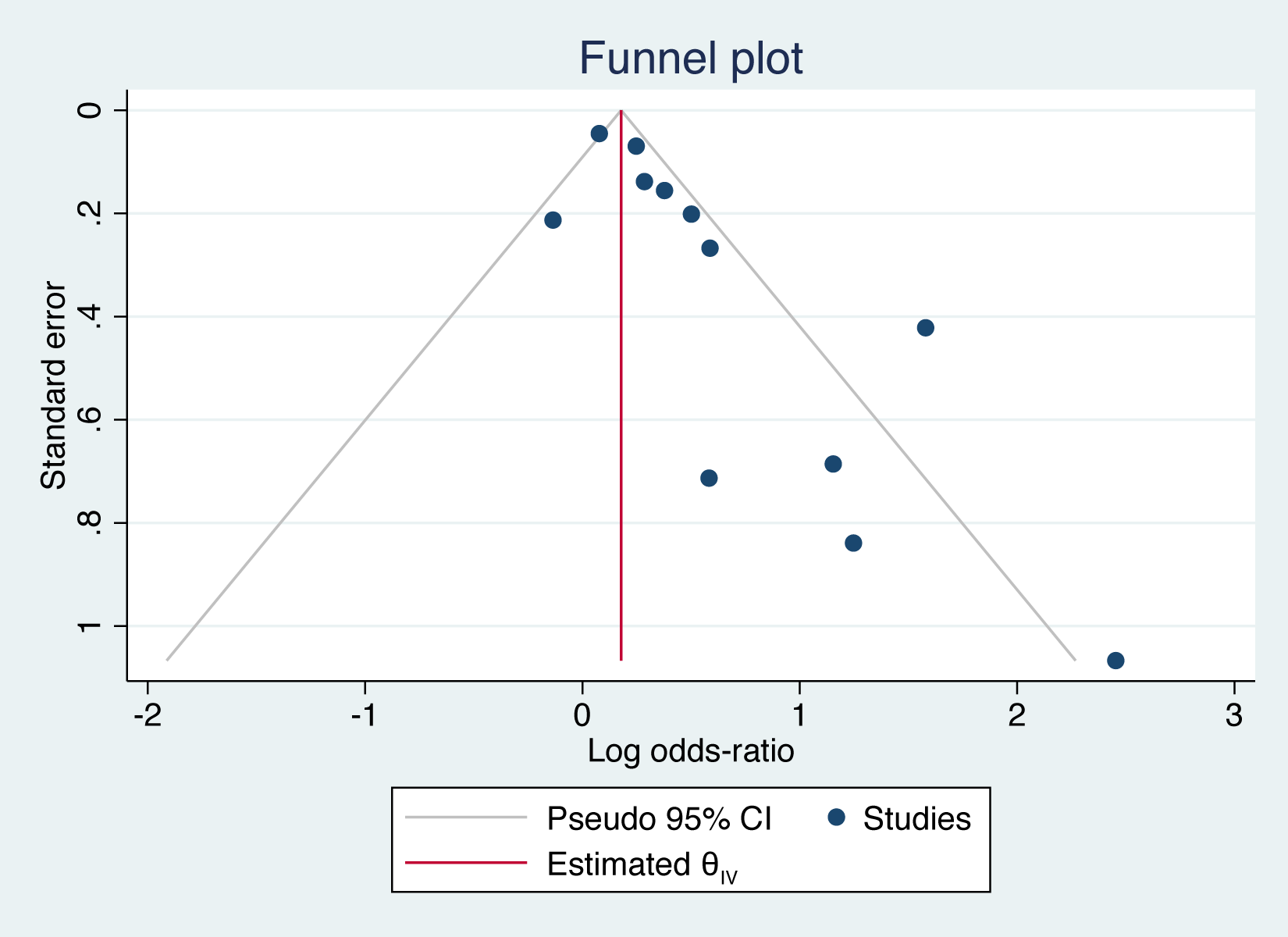

The main sources of heterogeneity for this meta-analysis are likely due to population size differences, disparities in detection methodology and recruitment Study design also evidently contributed to the I^2^ statistic, shown by the moderate to low heterogeneity within each design’s sub-analysis (Figure 3).

This result, when taken without correction for potential publication bias, shows a considerably raised risk when compared with other known CRC risk factors. With red-meat consumption at a risk ratio of 1.12 (95% CI 1.03 – 1.21),[18] and obesity at 1.19 (95% CI 1.11-1.29),[19] *H. pylori* infection proves to be a significant risk for CRC carcinogenesis.

The analyses discussed here showed evidence of publication bias. After running trim-and-fill, it was calculated that the addition of the 5 studies theoretically missing from the results would have resulted in a statistically insignificant link between *H. pylori* infection and CRC (Supplemental Material S7). Without these studies however, it is impossible to tell whether this is correct. Therefore, the result as it stands indicates; infection with *H. pylori* significantly increases the risk of CRC.

#### Limitations of included studies

There was a variation in the methods used to detect *H. pylori* infection. Many used detections of IgG through ELISA, others using tests such as urease and carbon breath tests. Using IgG detection, it is impossible to work out whether the infection is active or already eradicated. Both historic and ongoing infection may increase CRC risk, however for this hypothesis to be tested effectively, infection must be confirmed as either active or eradicated alongside IgG detection.

#### Limitations of this Meta-Analysis

As the initial aim of the paper was not to perform a meta-analysis on this subject, this search strategy was not fully comprehensive. An extra search into *H. pylori* alone may have returned more papers and provided additional data for the analysis.

The large degree of heterogeneity may be a limitation, potentially affecting the clinical significance of this meta-analysis and its results.

### Future research

As discussed, the case-control design is not sufficient to determine causality in this instance. This paper has identified the most frequently replicated key genera and species linked to CRC. However, due to the limitations of these study designs, it cannot be concluded that these bacteria raise the risk of developing CRC. To study this effectively, a large population prospective cohort study should be performed, with regular colonoscopy and microbiome sampling. This sampling should be a combination of faecal, swab and biopsy sampling. This would facilitate the representative characterisation of the microbiome profiles most commonly linked to CRC development, allowing a conclusion to be drawn as to whether there is any causative relationship between the human gut bacterial microbiome and CRC.

### Implications of this paper

To the researchers’ knowledge, this is the first systematic review of this type conducted into this topic. The results of this paper, combined with those of prospective cohort studies as suggested, may have implications in the personalised management of patients to reduce their risk of CRC. The potential implications of the use of probiotics to lower risk has not yet been fully investigated and requires serious attention. However, this paper has identified species that may be of interest in a study of this nature. If CRC associated bacteria can be reduced in abundance within the microbiome, and healthy associated bacteria increased, this may reduce an individual’s risk of developing CRC.

## CONCLUSION

The systematic review demonstrates the paucity and discrepancies amongst current research into the elevated risk bacteria has on CRC development. Although, considering these limitations, certain consistencies were available within the data extracted. There is a considerable link between the *Fusobacterium* genus and CRC, with a possible link suggested between bacterial presence of *Fusobacterium nucleatum* and *Bacteroides fragilis* and the increased CRC risk associated. Additionally, the meta-analysis suggested a significant association between *H. pylori* presence and CRC risk. However, both review and analysis require further expansion in methodology as well as an improvement in research papers looking to elucidate a more significant association. This assessment finds the need for a large-scale cohort study over a significant period instead of a case-control, in order to elucidate a potential carcinogenic relationship. This systematic review has documented all bacterial genera and species that were significantly enriched in CRC patients or controls in eligible papers. The hope is this will help guide future research into the role of the microbiome in CRC carcinogenesis, with the goal of more personalised management of CRC case prevention for patients in the future.

## Data Availability

All data relevant to the study are included in the article or uploaded as supplemental information

## Contributors

CAR: study design, search strategy, critical appraisal, data extraction, meta-analysis, manuscript writing; NAZ: study design, search strategy, critical appraisal, data extraction, meta-analysis, manuscript writing; VN: study design, search strategy, manuscript writing

CAR and NAZ contributed equally to this paper.

## Patient and public involvement

Patients and/or the public were not involved in any aspect of this research

## Data availability statement

All data relevant to the study are included in the article or uploaded as supplemental information

