## Supplemental Material S1 for "Gut Bacterial Microbiome Profiles Associated with Colorectal Cancer Risk: A Systematic Review and Meta-Analysis"

### EMBASE

1. colorectal cancer.mp. or exp colorectal cancer/

2. ((colo* or rectal) adj3 (cancer* or neoplas* or tumo?r* or malignanc* or carcino*)).mp. [mp=title, abstract, heading word, drug trade name, original title, device manufacturer, drug manufacturer, device trade name, keyword, floating subheading word, candidate term word]

3. 1 or 2

4. exp bacterium/

5. exp microbiome/ or Microbiome.mp.

6. Microb*.mp.

7. Intestinal microb*.mp. [mp=title, abstract, heading word, drug trade name, original title, device manufacturer, drug manufacturer, device trade name, keyword, floating subheading word, candidate term word]

8. Gastrointestinal microb*.mp. [mp=title, abstract, heading word, drug trade name, original title, device manufacturer, drug manufacturer, device trade name, keyword, floating subheading word, candidate term word]

9. Colonic microb*.mp. [mp=title, abstract, heading word, drug trade name, original title, device manufacturer, drug manufacturer, device trade name, keyword, floating subheading word, candidate term word]

10. F?ecal microb*.mp. [mp=title, abstract, heading word, drug trade name, original title, device manufacturer, drug manufacturer, device trade name, keyword, floating subheading word, candidate term word]

11. Gut microb*.mp. [mp=title, abstract, heading word, drug trade name, original title, device manufacturer, drug manufacturer, device trade name, keyword, floating subheading word, candidate term word]

12. Gut bacteria.mp. [mp=title, abstract, heading word, drug trade name, original title, device manufacturer, drug manufacturer, device trade name, keyword, floating subheading word, candidate term word]

13. Dysbiosis.mp. or exp dysbiosis/

14. 4 or 5 or 6 or 7 or 8 or 9 or 10 or 11 or 12 or 13

15. 3 and 14

16. exp case control study/ or case control stud*.mp.

17. Cohort stud*.mp. or exp Cohort Studies/

18. prospective stud*.mp.

19. 16 or 17 or 18

20. 15 and 19

### Medline

1. colorectal cancer.mp. or exp Colorectal Neoplasms/

2. ((colo* or rectal) adj3 (cancer* or neoplas* or tumo?r* or malignanc* or carcino*)).mp. [mp=title, abstract, original title, name of substance word, subject heading word, floating sub-heading word, keyword heading word, organism supplementary concept word, protocol supplementary concept word, rare disease supplementary concept word, unique identifier, synonyms]

3. 1 or 2

4. exp Bacteria/ or microb*.mp. or exp Microbiota/

5. Intestinal microb*.mp. [mp=title, abstract, original title, name of substance word, subject heading word, floating sub-heading word, keyword heading word, organism supplementary concept word, protocol supplementary concept word, rare disease supplementary concept word, unique identifier, synonyms]

6. Gastrointestinal microb*.mp. [mp=title, abstract, original title, name of substance word, subject heading word, floating sub-heading word, keyword heading word, organism supplementary concept word, protocol supplementary concept word, rare disease supplementary concept word, unique identifier, synonyms]

7. Colonic microb*.mp. [mp=title, abstract, original title, name of substance word, subject heading word, floating sub-heading word, keyword heading word, organism supplementary concept word, protocol supplementary concept word, rare disease supplementary concept word, unique identifier, synonyms]

8. F?ecal microb*.mp. [mp=title, abstract, original title, name of substance word, subject heading word, floating sub-heading word, keyword heading word, organism supplementary concept word, protocol supplementary concept word, rare disease supplementary concept word, unique identifier, synonyms]

9. Gut microb*.mp. [mp=title, abstract, original title, name of substance word, subject heading word, floating sub-heading word, keyword heading word, organism supplementary concept word, protocol supplementary concept word, rare disease supplementary concept word, unique identifier, synonyms]

10. Gut bacteria.mp. [mp=title, abstract, original title, name of substance word, subject heading word, floating sub-heading word, keyword heading word, organism supplementary concept word, protocol supplementary concept word, rare disease supplementary concept word, unique identifier, synonyms]

11. Dysbiosis.mp. or exp Dysbiosis/

12. 4 or 5 or 6 or 7 or 8 or 9 or 10 or 11

13. 3 and 12

14. exp Case-Control Studies/ or Case control stud*.mp.

15. exp Cohort Studies/ or Cohort stud*.mp.

16. Prospective stud*.mp.

17. 14 or 15 or 16

18. 13 and 17
