## Supplemental Material S2 for "Gut Bacterial Microbiome Profiles Associated with Colorectal Cancer Risk: A Systematic Review and Meta-Analysis"

*Details of studies specific to bacterial species*

| Author | Specific Bacteria Studied |
|---|---|
| **Tunsjø et al. (2019)** | *Fusobacterium nucleatum* |
| **Geravand et al. (2019)** | *Enterococcus faecalis* |
| **Suehiro et al. (2017)** | *Fusobacterium nucleatum* |
| **Liang et al. (2017)** | *Bacteroides clarus, Clostridium hathewayi, Fusobacterium nucleatum & Roseburia intestinalis* |
| **Zhou et al. (2016)** | *Bacteroides fragilis, Enterococcus faecalis, Fusobacterium spp. & Escherichia coli* |
| **Keenan et al. (2016)** | *Bacteroides fragilis* (Enterotoxigenic) |
| **Paritsky et al. (2015)** | *Streptococcus bovis* |
| **Magdy et al. (2015)** | *Escherichia coli* (Enteropathogenic) |
| **Boltin et al. (2015)** | *Streptococcus bovis* |
| **Kohoutova et al. (2015)** | *Escherichia coli* |
| **Potter et al. (1998)** | *Streptococcus bovis* |
