## Supplemental Material S3 for "Gut Bacterial Microbiome Profiles Associated with Colorectal Cancer Risk: A Systematic Review and Meta-Analysis"

| CASP Checklist | Section A | | | | | | Section B | Section C | |
|---|---|---|---|---|---|---|---|---|---|
| Paper | 1 | 2 | 3 | 4 | 5 | 6b | 9 | 10 | 11 |
| Liu (2020) | Yes | Yes | Yes | Yes | Yes | Yes | Yes | Yes | Yes |
| Zhang (2019) | Yes | Yes | Yes | Can't tell | Yes | Yes | Yes | No | Yes |
| Yachida (2019) | Yes | Yes | Can't tell | Can't tell | Can't tell | Yes | Yes | Can't tell | Yes |
| Tunsjo (2019) | Yes | Yes | Yes | No | Yes | Yes | Yes | No | Yes |
| Saito (2019) | Yes | Yes | Yes | No | Yes | Yes | Yes | No | Yes |
| Liu (2019) | Cohort study | | | | | | | | |
| Geravand (2019) | Yes | Yes | Can't tell | Can't tell | Yes | Yes | Yes | No | Yes |
| Butt (2019) | Yes | Yes | Yes | Yes | Can't tell | Yes | No | Yes | No |
| Zhang (2018) | Yes | Yes | No | No | Yes | Yes | Yes | No | Yes |
| Rezasoltani (2018) | Yes | Yes | Yes | Yes | Yes | Yes | Yes | Yes | Yes |
| Mori (2018) | Yes | Yes | No | No | Yes | Yes | Yes | No | Yes |
| Allali (2018) | Yes | Yes | Can't tell | Can't tell | Yes | Yes | Yes | Yes | Yes |
| Yu (2017) | Yes | Yes | Can't tell | Can't tell | Can't tell | Yes | Yes | Can't tell | Yes |
| Xu (2017) | Yes | Yes | Can't tell | Can't tell | Yes | Yes | Yes | Can't tell | Yes |
| Suehiro (2017) | Yes | Yes | Yes | Yes | Yes | Yes | Yes | Yes | No |
| Liang (2017) | Yes | Yes | Yes | No | Yes | Yes | Yes | Yes | Yes |
| Flemer (2017) | Yes | Yes | Yes | Can't tell | Yes | Yes | Yes | Can't tell | Yes |
| Zhou (2016) | Yes | Yes | Yes | Yes | Yes | Yes | Yes | Yes | Yes |
| Vogtmann (2016) | Yes | Yes | Can't tell | Can't tell | Can't tell | Yes | Yes | No | Yes |
| Thomas (2016) | Yes | Yes | Yes | No | Yes | Yes | Yes | Can't tell | Yes |
| Sinha (2016) | Yes | Yes | Can't tell | No | Can't tell | Yes | No | No | Yes |
| Keenan (2016) | Yes | Yes | Can't tell | Can't tell | Yes | Yes | Yes | No | Yes |
| Paritsky (2015) | Cohort study | | | | | | | | |
| Mira-Pascual (2015) | Yes | Yes | Yes | Yes | Yes | Yes | Yes | No | Yes |
| Magdy (2015) | Yes | Yes | Yes | Yes | Yes | Yes | Yes | No | Yes |
| Feng (2015) | Yes | Yes | Yes | Can't tell | Yes | Yes | Yes | Yes | Yes |
| Boltin (2015) | Cohort Study | | | | | | | | |
| Zeller (2014) | Yes | Yes | No | No | Yes | No | Can't tell | Can't tell | Can't tell |
| Shmuely (2014) | Cross-Sectional Study | | | | | | | | |
| Selgrad (2014) | Cross-Sectional Study | | | | | | | | |
| Kohoutova (2014) | Yes | Yes | Yes | No | Yes | Yes | Yes | Yes | Yes |
| Hsu (2014) | Cohort Study | | | | | | | | |
| Flanagan (2014) | Yes | No | No | No | Yes | No | No | No | Yes |
| Weir (2013) | Yes | Yes | Can't tell | Can't tell | Yes | Yes | Yes | Yes | Yes |
| Ohigashi (2013) | Yes | No | Yes | No | Can't tell | Yes | Yes | No | Yes |
| Nam (2013) | Yes | Yes | Yes | Yes | Yes | Yes | Yes | Can't tell | Yes |
| Ahn (2013) | Yes | Yes | No | No | Yes | Yes | Yes | Yes | Yes |
| Zhang (2012) | Yes | Yes | Yes | Yes | Yes | Yes | Yes | Yes | Yes |
| Chen (2012) | Yes | No | Yes | Can't tell | Yes | No | Yes | Yes | Yes |
| Sobhani (2011) | Yes | Yes | Yes | No | Yes | Yes | Yes | Yes | Can't tell |
| Abbass (2011) | Cross-sectional study | | | | | | | | |
| Zumkeller (2007) | Yes | Yes | Yes | Yes | Yes | Yes | Yes | Yes | Yes |
| Jones (2007) | Yes | Yes | Yes | No | Yes | Yes | Yes | Yes | Yes |
| Fujimori (2005) | Yes | Yes | Yes | No | Yes | No | No | No | Yes |
| Siddheshwar (2001) | Yes | Yes | Yes | No | Yes | Yes | Yes | No | Yes |
| Potter (1998) | Yes | Yes | Yes | No | No | No | Yes | Yes | Yes |

| Key | | |
|---|---|---|
| Accepted Studies | | Yes |
| Rejected Studies | | Can't tell |
| | | No |

| Newcastle-Ottawa | Selection (max. 5 stars) | | | | Comparability | Outcome (max. 3 stars) | |
|---|---|---|---|---|---|---|---|
| Paper | 1 | 2 | 3 | 3 | 1 | 1 | 2 |
| Shmuely et al. (2014) | * | * | | ** | * | ** | * |
| Selgrad et al. (2014) | * | * | | ** | * | ** | * |
| Abbass et al. (2011) | * | * | | ** | * | ** | * |

| CASP checklist | Section A | | | | | | | | Section B | Section C | | |
|---|---|---|---|---|---|---|---|---|---|---|---|---|
| Paper | 1 | 2 | 3 | 4 | 5a | 5b | 6a | 6b | 9 | 10 | 11 | 12 |
| Liu 2019 | | | | | | | | | | | | |
| Paritsky 2015 | | | | | | | | | | | | |
| Boltin et al. (2015) | | | | | | | | | | | | |
| Hsu (2014) | | | | | | | | | | | | |
