## Supplemental Material S4 for "Gut Bacterial Microbiome Profiles Associated with Colorectal Cancer Risk: A Systematic Review and Meta-Analysis"

| Author | Pop. | N | Sampling | Detection Method | Taxa Enriched in CRC | Taxa Enriched in Control |
|---|---|---|---|---|---|---|
| Liu et al. 2020 [32] | China | 147 | Faecal | Illumina | **Proteobacteria, Spirochaetes, Synergistetes** | **Firmicutes** |
| Zhang et al. 2019 [33] | China | 23 | Biopsy | Illumina | *Devosia* | *Blautia producta, Eubacterium, Klebsiella, Leptotrichia, Phascolarctobacterium, Prevotella copri & stercorea* |
| Yachida et al. 2019 [34] | Japan | 616 | Faecal | WGSS | *Bilophila wadsworthia, Collinsella aerofaciens, Desulfovibrio longreachensis* & vietnamensis, Dorea longicatena, Fusobacterium nucleatum, Gemella morbillorum, Lactobacillus sanfrancisensis, Parvimonas micra, Peptostreptococcus anaerobius & stomatis, Phascolarctobacterium succinatutens*, Porphyromonas ueonis, Selenomas sputigena, Solobacterium moorei, Streptococcus angionus* | *Eubacterium eligens, Lachnospira multipara* |
| Tunsjø et al. 2018 [35] | Norway | 77 | Faecal & Biopsy | qPCR | *Fusobacterium nucleatum* | |
| Saito et al. 2019 [12] | Japan | 81 | Colonoscopy Aspirate | Illumina | *Actinobacillus, Actinomyces, Fusobacterium (varium), Parvimonas, Peptostreptococcus* | *Fusobacterium peridonticum, Megamonas, Sphingobium* |
| Geravand et al. 2019 [36] | Iran | 77 | Faecal | qPCR | *Enterococcus faecalis* | |
| Zhang et al. 2018 [37] | China | 348 | Faecal | Illumina | *Campylobacter rectus, Clostridium lactatifermentans, Clostridium scindens, Clostridium symbiosum, Dialister pneumosintes, Eggerthella lenta, Eisenbergiella tayi, Fusobacterium nucleatum, Gemella morbillorum, Parvimonas micra, Peptostreptococcus stomatis, Porphyromonas asaccharolytica, Rumminococcus torques, Solobacterium moorei* | *Blautia faecis, Coprococcus comes, Eubacterium desmolans, Eubacterium eligens, Eubacterium hadrum, Eubacterium hallii, Fusicatenibacter saccharivorans, Roseburia faecis, Ruminococcus lactaris, Streptococcus salivarius* |
| Rezasoltani et al. 2018 [38] | Iran | 93 | Faecal | qPCR | *Enterococcus faecalis, Bacteroides fragilis,* Enterotoxigenic *Bacteroides fragilis, Fusobacterium* | |

| Author | Pop. | N | Sampling | Detection Method | Taxa Enriched in CRC | Taxa Enriched in Control |
|---|---|---|---|---|---|---|
| | | | | | *nucleatum, Porphyromonas (gingivalis), Streptococcus bovis* | |
| Mori et al. 2018 [39] | Italy | 65⁺ | Faecal | Illumina | *Escherichia, Shigella, Sutterella* | *Anaerostipes* |
| Allali et al. 2018 [40] | Morocco | 23 | Faecal | Illumina | *Akkermansia (municiphilia), Bacteroides fragilis, Bilophila, Butyricimonas, Christensenella, Clostridium, Collinsella aerofaciens, Dehalobacterium, Eubacterium (biforme), Fusobacterium, Oscillospira, Oxalobacter (formigenes), Parabacteroides, Peptostreptococcus, Porphyromonas, Ruminococcus, Selenomas* | *Faecalibacterium prausnitzii, Megamonas, Prevotella copri, Prevotella stercorea* |
| Yu et al. 2017 [41] | Hong Kong & Denmark | 324 | Faecal | Illumina | *Bacteroides fragilis, Fusobacterium (nucleatum), Gemella (morbillorum), Parvimonas (micra), Peptostreptococcus (stomatis), Solobacterium (moorei)* | *Eubacterium (ventriosum)* |
| Xu and Jiang 2017 [42] | China | 160 | Previous Data | | *Campylobacter, Dialister, Fusobacterium, Lactobacillus, Leptotrichia, Mogibacterium, Parvimonas, Peptostreptococcus* | *Acidomonas, Blautia, Escherichia, Faecalibacterium, Pseudomonas, Sphingomonas* |
| Suehiro et al. 2017 [43] | Japan | 109 | Faecal | Droplet PCR | *Fusobacterium nucleatum* | |
| Liang et al. 2017 [44] | China & Hong Kong | 536 | Faecal | qPCR | *Fusobacterium nucleatum, Clostridium hathewayi* | *Bacteroides clarus, Roseburia intestinalis* |
| Flemer et al. 2017 [45] | Ireland | 147 | Biopsy | Illumina | *Bacteroides, Fusobacterium, Oscillibacter, Roseburia, Ruminococcus.* | *Coprococcus, Lachnospiraceae* |
| Zhou et al. 2016 [46] | China | 145 | Biopsy | qPCR | *Enterococcus (faecalis), Fusobacterium* | |
| Vogtmann et al. 2016 [47] | USA | 104 | Lyophilised Faeces | WGSS | *Fusobacterium, Porphyromonas* | |

| Author | Pop. | N | Sampling | Detection Method | Taxa Enriched in CRC | Taxa Enriched in Control |
|---|---|---|---|---|---|---|
| Thomas et al. 2016 [48] | Brazil | 36 | Biopsy | QIIME | *Bacteroides (fragilis, uniformis), Bilophila, Desulfovibrio, Fusobacterium, Odoribacter, Parabacteroides, Phascolarctobacterium* | *Acinetobacter, Alcaligenes faecalis, Bacillus (cereus), Escherichia, Lactobacillus (delbruecki), Prevotella melaninogenica, Pseudomonas* |
| Sinha et al. 2016 [49] | USA | 150 | Lyophilised faeces | QIIME | *Fusobacterium, Porphyromonas* | |
| Keenan et al. 2016 [50] | New Zealand | 142 | Faecal | qPCR | Enterotoxigenic *Bacteroides fragilis* | |
| Paritsky et al. 2015 [51] | Israel | 203 | Colonoscopy Aspirate | Culturing | *Streptococcus bovis* | |
| Mira-Pascual et al. 2015 [52] | Spain | 28 | Faecal & Biopsy | Pyro | **Enterobacteriaceae**, *Blautia coccoides* | |
| Magdy et al. 2015 [53] | Egypt | 461 | Biopsy | Antiserum | Enteropathogenic *Escherichia coli* | |
| Feng et al. 2015 [54] | Austria | 138 | Faeces | Illumina | *Acidaminococcus (intestini), Alistipes (finegoldii, putredinis), Bacteroides (caccae, dorei, eggerthii, massilliensis, ovatus, vulgatus, xylanisolvens), Bilophila (wadsworthia), Burkholderia, Clostridium (symbiosum), Escherichia (coli), Lachnospiracae, Odoribacter (splanchius), Parabacteroides (distasonis, merdae), Paraprevotella (clara), Sutterella (wadsworthensis), Veilonella (atypica)* | *Actinomyces (viscosus), Bifidobacterium (animalis), Clostridium, Streptococcus (mutans, thermophilus)* |
| Boltin et al. 2015 [11] | Israel | 118 | Colonoscopy Aspirate & Biopsy | Culturing | No significance found | No significance found |
| Kohoutova et al. 2014 [55] | Czech Republic | 80 | Biopsy | Culturing & Primers | *Escherichia coli* | |

| Author | Pop. | N | Sampling | Detection Method | Taxa Enriched in CRC | Taxa Enriched in Control |
|---|---|---|---|---|---|---|
| Weir et al. 2013 [56] | USA | 21 | Faecal | Pyro | *Acidaminobacter (unspecified), Akkermansia (muciniphilia), Citrobacter (farmeri), Phascolarctobacterium (unspecified)* | *Bacteroides (capillosus, finegoldii, intestinalis), Dialister (invisus, pneumosintes), Dorea (formicigenerans), Lachnobacterium (bovis), Lachnospira (pectinoschiza), Megamonas (hypermegale), Prevotella (copri, oris), Pseudobutyrovibrio (ruminis), Ruminococcus (albus, obeum)* |
| Ahn et al. 2013 [57] | USA | 141 | Lyophilised Faeces | QIIME | *Anaerovorax, Atopobium, Fusobacterium, Megasphaera, Peptostreptococcus, Porphyromonas, Selenomas* | *Coprococcus, Ruminococcus* |
| Chen et al. 2012 [17] | China | 102 | Swab, Faecal & Biopsy | Pyro | *Anaerococcus*[!], *Anaerotruncus*[!], *Catonella*[$], *Collinsella*[!], *Desulfovibrio*[!], *Eubacterium*[!], *Filifactor*[$], *Fusobacterium*[$], *Gemella*[$], *Klebsiella*[$], *Mogibacterium*[$!], *Paraprevotella*[!], *Peptostreptococcus*[$!], *Slackia*[!], *Porphyromonas*[$], *Selenomas*[$] | *Anaerostipes, Bifidobacterium, Blautia, Catenibacterium, Faecalibacterium, Gardnerella, Lachnospira* |
| Sobhani et al. 2011 [58] | France | 179 | Faecal | Pyro | *Bacteroides/Prevotella* group | |
| Potter et al. 1998 [59] | UK | 42 | Faecal & Biopsy | Culturing | No significance found | No significance found |

Where a species is in brackets next to its corresponding genus, denotes that both genus and species are statistically significant. If species not in brackets, only the species is significant.

[+] = article contains more patients that had undergone chemotherapy, these are excluded from numbers and results.

[$]= faecal sample, [!] = swab sample, WGSS = Whole Genome Shotgun Sequencing, Pyro = Pyrosequencing. Where genus level is not available or scarce, family taxonomic level and above was included and listed in bold.
