## Supplemental Material S6 for "Gut Bacterial Microbiome Profiles Associated with Colorectal Cancer Risk: A Systematic Review and Meta-Analysis"

```
Regression-based Egger test for small-study effects
Random-effects model
Method: REML

H0: beta1 = 0; no small-study effects
            beta1 =       1.75
      SE of beta1 =      0.441
                z =       3.98
       Prob > |z| =     0.0001
```
