## Supplemental Material S7 for "Gut Bacterial Microbiome Profiles Associated with Colorectal Cancer Risk: A Systematic Review and Meta-Analysis"

```
. meta trimfill, eform funnel(contours(1 5 10))

  Effect-size label:  Log Odds-Ratio
        Effect size:  _meta_es
          Std. Err.:  _meta_se

Nonparametric trim-and-fill analysis of publication bias
Linear estimator, imputing on the left

Iteration                              Number of studies =     17
  Model: Random-effects                         observed =     12
 Method: REML                                   imputed =      5

Pooling
  Model: Random-effects
 Method: REML
```

| Studies | Odds Ratio | [95% Conf. | Interval] |
|---|---|---|---|
| Observed | 1.485 | 1.187 | 1.858 |
| Observed + Imputed | 1.260 | 0.928 | 1.711 |
